# The DIAMONDS intervention to support self-management of type 2 diabetes in people with severe mental illness: study protocol for a single-group feasibility study

**DOI:** 10.1101/2021.12.05.21267169

**Authors:** Jennifer V E Brown, Ramzi Ajjan, Sarah Alderson, Jan R Böhnke, Claire Carswell, Patrick Doherty, Keith Double, Simon Gilbody, Michelle Hadjiconstantinou, Catherine Hewitt, Richard I G Holt, Rowena Jacobs, Vicki Johnson, Ian Kellar, David Osborn, Steve Parrott, David Shiers, Johanna Taylor, Jacqui Troughton, Judith Watson, Najma Siddiqi, Peter A Coventry, on behalf of the DIAMONDS research team

**Author notes:** corresponding author: Jennifer V E Brown, Mental Health & Addiction Research Group, Department of Health Sciences, University of York, York, YO10 5DD, UK.

## Abstract

**Introduction:** The DIAMONDS programme aims to evaluate a novel supported diabetes self-management intervention for people with severe mental illness (the “DIAMONDS intervention”). The purpose of this study is to test the feasibility of intervention delivery and data collection procedures to inform a definitive randomised controlled trial (RCT).

**Methods:** Adults aged 18 years or over with a diagnosis of type 2 diabetes and severe mental illness (schizophrenia, schizoaffective disorder, or bipolar disorder) will be eligible for inclusion. Individuals with other types of diabetes or non-psychotic mental illness and those lacking capacity to consent will not be eligible. Participants will be recruited from NHS mental health trusts and general practices across the North of England. All participants will receive the DIAMONDS intervention: weekly one-to-one sessions with a trained facilitator (“DIAMONDS Coach”) to support goal setting, action planning, and diabetes education; ongoing self-management supported by a paper-based workbook and optional digital application (app); and monthly peer-support group sessions with other participants. The primary outcomes are: 1. Recruitment rate, measured as proportion of the recruitment target (N=30) achieved at 5 months from start of recruitment, 2. Attrition measured as the proportion of missing outcomes data at the end of the recruitment period (5 months from start of recruitment) for physiological and self-reported data items, 3. Intervention delivery rate recorded as the proportion of planned sessions delivered (measured by the number of completed intervention session logs per participant within 15 weeks of the first intervention session). Secondary outcomes include completeness of data collection at baseline and of process evaluation data at follow-up as well as the feasibility and acceptability of the intervention and of wearing a blinded continuous glucose monitoring device. An intervention fidelity framework will also be developed. Recruitment started in July 2021. The study was prospectively registered: ISRCTN15328700 (12^th^ March 2021).

**Discussion:** The results of this feasibility study will inform the refinement of the content and delivery of the DIAMONDS intervention, as well as research procedures, including recruitment and data collection, in preparation for the main DIAMONDS RCT.

## 1 Introduction

People with severe mental illness (SMI; i.e. long-term mental illnesses such as schizophrenia, schizoaffective disorder, and bipolar disorder)[1] experience higher rates of physical illness and poorer health outcomes than the general population. People with SMI die 15-20 years earlier than the general population,[2-5] mainly from comorbid long-term conditions (LTCs).[6-8] The provision of clinically and cost-effective healthcare for people with combinations of mental and physical illness is recognised as challenging.[9] Symptoms associated with co-existing mental and physical conditions and treatment regimens may interact antagonistically and exacerbate disease and treatment burden.[10]

Diabetes is two to three times more common in people with SMI than in the general population,[5, 11] and is associated with poorer outcomes than for individuals with diabetes alone.[6-8] Over 99% of diabetes care falls to self-management.[12] Appropriate self-management in diabetes (in common with other LTCs) is fundamental to improving clinical outcomes in this population.[13-15] Self-management refers to the skills, practices, and behaviours that a person engages in to protect and promote their health. Diabetes self-management activities include: consuming a healthy diet; increasing physical activity; smoking cessation; monitoring glycaemic levels; preventing complications; and taking medicines as prescribed.[16, 17] “Self-management education” is key to supporting self-management.[15, 18, 19] In England, diabetes self-management education programmes are recommended for people with recently diagnosed type 2 diabetes and their family members or supporters.[15] Such programmes typically include educational and behavioural elements to increase knowledge, skills, and ability for self-management,[20] and target healthy diet, exercise, smoking cessation, appropriate self-monitoring, and medication taking.[15, 21] Self-management education programmes for the general population with diabetes have been found to be both clinically worthwhile and cost-effective.[12, 20, 22-25]

For people with SMI and diabetes, self-management support is rarely offered, although reliable data on this are difficult to obtain.[26] Moreover, the effectiveness of diabetes self-management programmes for people with SMI is largely unknown as research typically excludes them.[27-29] SMI is characterised by disturbances of thought, perception, affect, and motivation,[30, 31] which may adversely influence self-efficacy, literacy, lifestyle, behaviour, and family life.[32-35] Diabetes self-management programmes designed for the general population do not address these important barriers.[36-39]

The DIAMONDS programme aims to develop and evaluate a diabetes self-management intervention for people with SMI and type 2 diabetes.[40] The goal of the intervention is to support people to engage in type 2 diabetes self-management behaviours, alongside managing mental health comorbidities in order to improve glycaemic levels.

We have developed a blended-delivery self-management intervention that is based on the Theoretical Domains Framework and its extension, the Mechanism of Action (MoA) framework. [41, 42] MoAs are the processes through which behaviour change techniques (BCTs) influence behaviour. These processes can relate to characteristics of the individual or characteristics of the wider social and physical environment. [42] This process involved systematically reviewing the literature with a focus on living with SMI and LTCs to identify MoAs that underpin candidate BCTs.[43-45]

Additionally, we conducted in-depth semi-structured interviews to better understand the lived experience of people with SMI and LTCs and their carers. Health professionals supporting people with SMI were also interviewed (DIAMONDS Quest) to ascertain barriers and facilitators to optimal diabetes care for this population. Findings from the qualitative interviews will be published separately. These qualitative data were integrated with the review findings to inform a consensus exercise to identify which MoA-BCT links and modes of delivery offered the most potential and utility to modify self-management behaviours in people with SMI and LTCs.

We then worked in partnership with people with SMI and diabetes, along with their family members/friends, as well as healthcare staff who support people with SMI and diabetes, to co-design prototypes of the diabetes self-management intervention. [46-49] Preliminary user testing of these prototypes has helped to establish acceptability and functionality of the intervention in the DIAMONDS Co-design study. Details of the development process and outcomes will also be reported separately; the intervention components are described in the methods section.

Before we can definitively test clinical and cost-effectiveness of the DIAMONDS intervention, we first need to test the feasibility and acceptability of study processes and the means to deliver the intervention.

### 1.1 Objectives

The objectives of the feasibility study are to:

1. Test the feasibility of procedures for recruitment and retention of participants.
2. Test the feasibility of quantitative and qualitative data collection.
3. Undertake an evaluation of the acceptability and feasibility of the DIAMONDS intervention.
4. Undertake an exploratory economic evaluation.
5. Undertake an exploratory evaluation of the acceptability and feasibility of continuous glucose monitors among individuals with type 2 diabetes and SMI.
6. Develop an intervention fidelity framework for use in a future RCT.

## 2 Material and methods

This study protocol is reported in line with the SPIRIT checklist for trial protocols which can be found in Appendix A. Ethical approval was obtained from the Research Ethics Committee Leeds West (reference: 21/YH/0059).

This is a protocol for a single-group feasibility study which incorporates a mixed-methods process evaluation. The study setting will include six mental health trusts (secondary care) and approximately ten general practices (primary care) in the North of England.

### 2.1 Study population

#### Inclusion criteria

The target population will be adults (aged 18 years or older) with SMI (schizophrenia, bipolar disorder, schizoaffective disorder, psychosis not otherwise specified) and type 2 diabetes (insulin and non-insulin treated). For participants recruited via health services, a diagnosis of SMI will be confirmed by specialist psychiatric services or by a general practitioner (GP) and be documented in the patient’s medical records in general practice or secondary care. The diagnosis of type 2 diabetes needs to be of at least three months’ duration and documented in the medical records. For participants who are recruited from third sector and mental health service user groups, self-reported type 2 diabetes will be confirmed from primary care medical records.

#### Exclusion criteria

People who have cognitive impairment and those with a diagnosis of gestational diabetes, type 1 diabetes, diabetes due to a specific genetic defect or secondary to pancreatitis or endocrine conditions will be excluded. We will also exclude patients who lack capacity to participate in the study, guided by the 2005 Mental Capacity Act.

### 2.2 Sample size

We aim to recruit 30 participants with SMI and type 2 diabetes. This sample size is sufficient to collect feasibility data on the specified outcomes and in line with the median sample size used in feasibility studies funded by NIHR.[50] In addition, approximately seven informal carers and up to ten healthcare professionals trained to deliver the DIAMONDS intervention (“DIAMONDS Coaches”) will be recruited to take part in qualitative interviews.

### 2.3 Service user recruitment and consent procedures

Recruitment of participants will employ methods successfully deployed in the SCIMITAR+[51], STEPWISE[52], and PRIMROSE[53] trials and use a staged consent procedure. All participant-facing documents were produced in collaboration with DIAMONDS Voice, our service user and carer group. Eligible patients will first receive a brief study information leaflet which will be followed by a study information pack containing an invitation letter to join the study, a participant information sheet, and consent form. Potential participants will be contacted a few days later to discuss the study and to arrange a face-to-face or virtual meeting where they will be given a further opportunity to ask questions about the study. If the individual is willing to join the study at this point, informed consent will be taken. A sample consent form can be found in Appendix B.

### 2.4 Recruitment sources

#### GP database screening

General practices will be asked to consult their SMI and LTC Quality and Outcomes Framework registers to screen for potentially eligible patients.[54] GPs at participating practices will check the lists produced by the database search to confirm eligibility. Study information documents will be sent from these organisations, following the staged process as described previously. Consent-to-contact (CTC) will be obtained from patients approached in this way prior to study information being sent out by the research team.

#### Primary care referral following annual health check

A brief study information leaflet and CTC form will be given to interested and potentially eligible patients during their health check. Once CTC has been obtained, the consent process as described previously will be followed.

#### Community mental health teams

Authorised staff at participating sites will run searches in databases held in secondary care and screen community mental health team caseloads for potentially eligible patients. Individuals identified will be approached using the processes as described previously.

#### Identification from previous studies and research cohorts

We will also explore the option to use records from previous related studies and existing research cohorts to contact potentially eligible patients who have given consent to be contacted about other research. We will send a brief study information leaflet and a CTC form to the interested individual to complete and return to the study team. Before sending out the study information pack, the study team will use a diabetes screening question over the telephone with all individuals who return the CTC form.

#### Recruitment from third sector and service user groups

We will further aim to recruit from relevant local third sector organisations and service user groups. People who are interested in taking part in the study will be directed to the person in the organisation/service supporting the study, or the DIAMONDS study team. As detailed earlier, they will be provided with a brief study information leaflet and will be asked to complete and return the CTC form.

Regardless of the route of recruitment, potential participants will have a further opportunity to clarify any points they did not understand and ask any questions when they meet a member of the Research & Development (R&D) team at the participating NHS site who will be responsible for taking consent and completing baseline measures. Potential participants identified outside of secondary care will also be consented by a member of the R&D team of the NHS trust whose care they are under. Only those under the care of a participating trust will be eligible. It will be emphasised that the participant may withdraw their consent to participate at any time, without having to provide a reason, and without it affecting their usual care or benefits to which they are entitled. The participant will also be informed that by consenting, they agree to their GP being made aware of their participation in the study and their medical records may be inspected by the study team. The person taking consent will have training and competence in assessing capacity and taking consent in people with SMI. Written informed consent will be obtained with both the participant and the researcher signing and dating the consent forms prior to enrolment. However, appropriate procedures for taking verbal consent over the telephone will be in place, where an in-person meeting is not possible. An overview of the study management and governance structures can be found in Appendix C.

### 2.5 The DIAMONDS intervention

The intervention content is described using the Template for Intervention Description and Replication (TIDieR) checklist (Appendix D).[55] Intervention materials cannot be offered open access at this stage because they have not undergone definitive evaluation yet and may be developed further in preparation for the main trial. All participants will receive the DIAMONDS intervention and access to standard care will continue as usual. The DIAMONDS intervention is a tailored self-management support programme to help people with type 2 diabetes and SMI self-manage their diabetes through:

- Increasing knowledge and skills for type 2 diabetes self-management.
- Providing support to increase physical activity levels and make healthier food choices.
- Identifying and addressing barriers to taking medications.
- Identifying and addressing sleep problems.
- Supporting participants to manage their diabetes within the context of fluctuating and low mood.
- Facilitating peer support.

The intervention will be delivered by a trained facilitator (the “DIAMONDS Coach”), over 16 weeks, using a combination of individual weekly sessions and daily use of a paper-based workbook (the “DIAMONDS Workbook”) which will be supported by daily use of an optional digital app (“Change One Thing”), as well as optional monthly group sessions (see Figure 1).

**Figure 1:**
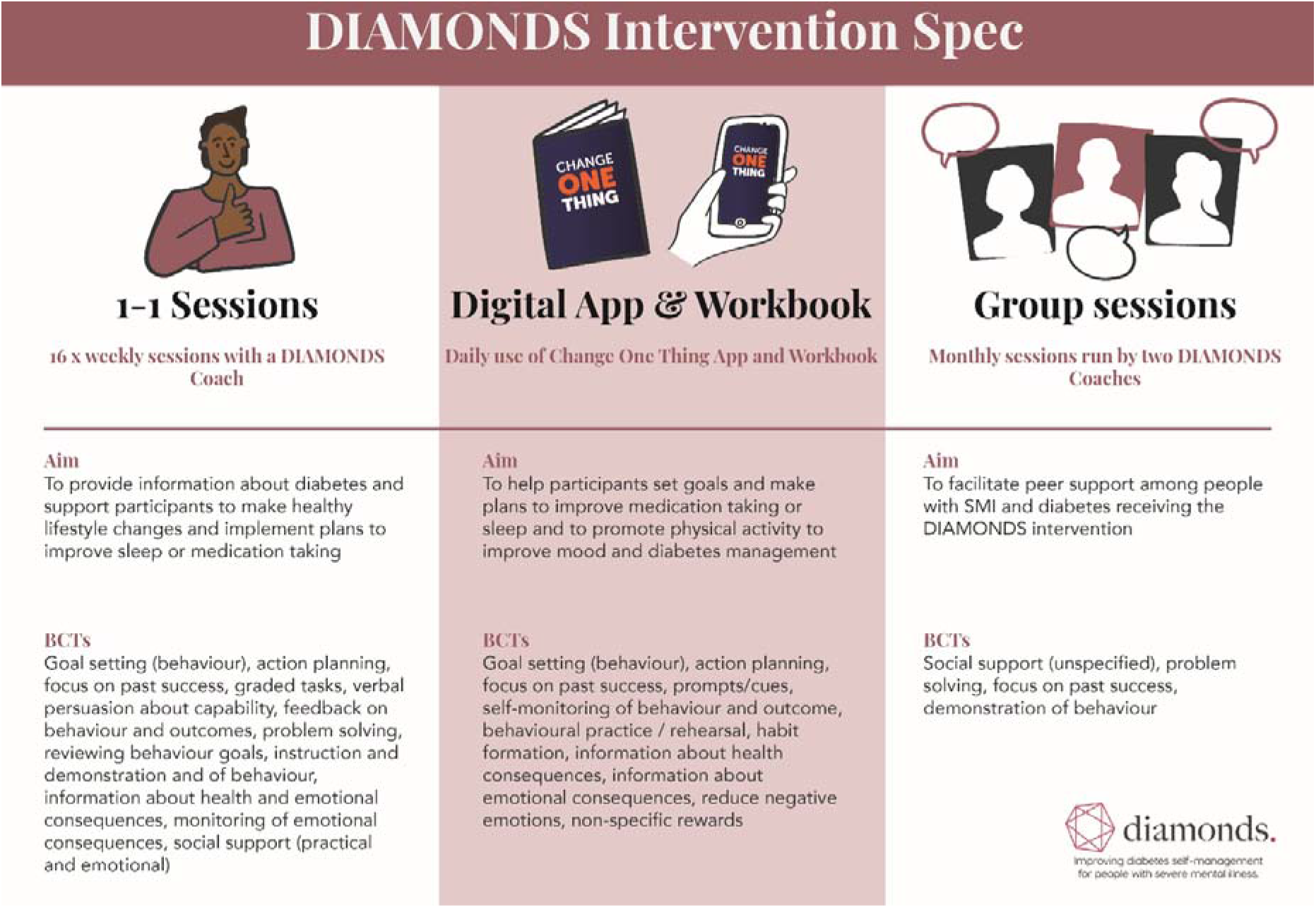
The DIAMONDS Intervention.

Using a specially developed and standardised training programme, we will train healthcare staff with experience of working in mental health services to become DIAMONDS Coaches. The training will be delivered by means of interactive workshops and supported by a training handbook and online videos and resources. All Coaches will receive and be instructed to use a Coach Manual to deliver the intervention. The Coach training materials and the Manual will cover the philosophy underpinning DIAMONDS, key coach behaviours, facilitation skills, and BCTs required to deliver the DIAMONDS intervention. Self-reflection will be encouraged and mentorship from the training team will also be provided.

Participants will be offered up to 16 individual weekly sessions with a DIAMONDS Coach. The first session will last between 60 and 90 minutes and follow-up sessions will each last between 30 and 60 minutes depending on the needs and preferences of the participant. All sessions will ideally be delivered face-to-face; however, they may be delivered by telephone or video call if participants express a preference for this and/or if ongoing COVID-19 restrictions necessitate remote delivery. In partnership with stakeholders, we decided to test a version of the intervention that allows us to work safely and take precautions to manage the risk of COVID-19 rather than redesigning it to be fully remote.

The main aim of the weekly sessions is to set goals and make plans to improve sleep, medication taking, or another area of self-management chosen by the participant in partnership with their Coach. In addition, the sessions aim to provide information about diabetes, and to support participants to increase physical activity levels and make other healthy lifestyle changes. Participants will be encouraged to engage with the intervention in between the weekly sessions. This process will be supported by the DIAMONDS Workbook and the Change One Thing app.

The intervention endpoint will be at 15 weeks after the participant’s first session with their DIAMONDS Coach regardless of the number of sessions attended by the participant. Session content will not be sequential but will instead be tailored to the participant’s needs; “missed” sessions will not necessarily mean that the participant misses out on intervention content. Participants will be able to continue engaging with intervention content on their own after follow-up process data are collected. From the outset, participants will be informed that sessions will stop 15 weeks after their first session. During the last month, the Coach will support participants to set longer-term goals and action plans for self-management and help them to access appropriate and existing support to implement these.

Where possible, monthly group sessions will be provided by two DIAMONDS Coaches (in a community-based venue, e.g. local town hall, community hub). Sessions will be held in the afternoons for 90 minutes. A healthy lunch will be provided at the start of each group session. Sessions will be attended by six to ten participants. The aim of the sessions is to facilitate peer-to-peer support among people with SMI and type 2 diabetes.

### 2.6 Service-user and carer involvement

Since the start of the DIAMONDS research programme in 2015, we have closely and continually collaborated with DIAMONDS Voice, a service-user and carer group dedicated to supporting this work. The group consists of approximately ten adults and includes members with severe mental illness as well as family carers. DIAMONDS Voice members have contributed critically to the intervention content as well as the development of the intervention materials (app and workbook). For this feasibility study, they reviewed all participant-facing documentation, including consent forms, invitation letters, and questionnaires, and were consulted about the acceptability of taking blood and undertaking measurements of their physical health.

### 2.7 Objectives 1 and 2: Testing the feasibility of study procedures

Outcomes for objective 1 (feasibility of recruitment and attrition):

a. Recruitment rate, measured as proportion of the recruitment target (N=30) achieved at 5 months from start of recruitment.
b. Attrition measured as the proportion of missing outcomes data at the end of the recruitment period (5 months from start of recruitment) for physiological and self-reported data items.
c. Intervention delivery rate recorded as the proportion of planned sessions delivered (measured by the number of completed intervention session logs per participant within 15 weeks of the first intervention session).

Outcomes for objective 2 (feasibility of data collection):

Additionally, the feasibility of quantitative data collection methods will be tested by collecting all planned primary and secondary outcomes for the main RCT as specified in table 1.

**Table 1:** Summary of data collection to be tested in the feasibility study.

| **Data item** | **Data collection method** | **Baseline** | **Follow-up** |
| --- | --- | --- | --- |
| HbA1c | Measured by study team | x | - |
| *Sociodemographics* |  |  |  |
| Age | Self-report | x | - |
| Sex | Self-report | x | - |
| Ethnicity | Self-report | x | - |
| Index of Multiple Deprivation | Determined by study team based on participant's postcode | x | - |
| *Physical health status* |  |  |  |
| Date diagnosed with diabetes | Primary care records | x | - |
| Height | Measured by study team | x | - |
| Weight | Measured by study team | x | - |
| BMI (calculated from height and weight) | Calculated by study team | x | - |
| Waist circumference | Measured by study team | x | - |
| Blood pressure | Measured by study team | x | - |
| Haemoglobin<sup>^</sup> | Measured by study team | - | - |
| Cholesterol<sup>^</sup> | Measured by study team | - | - |
| Medical co-morbidities | Primary care records | x | - |
| *Smoking status* | Self-report | x | - |
| *Mental health* |  |  |  |
| Type of SMI | Primary care records | x | - |
| Date diagnosed with SMI | Primary care records | x | - |
| Brief Psychiatric Rating Scale (BPRS) | Self-report | x | - |
| Patient Health Questionnaire-9 (PHQ-9) | Self-report | x | - |
| *Diabetes measures* |  |  |  |
| Diabetes microvascular and macrovascular complications | Medical record | x | - |
| Diabetes distress | Self-report | x | - |
| Summary of Diabetes Self-Care Activities | Self-report | x | - |
| *Physical activity* |  |  |  |
| International Physical Activity Questionnaire (IPAQ) | Self-report | x | - |
| Physical activity | Wrist-worn accelerometer | x | - |

| *Health economic outcomes* |  |  |  |
| --- | --- | --- | --- |
| Health-related quality of life (EQ-5D-5L) | Self-report | x | - |
| Health resource use | Self-report & Medical records | x | - |
| Medication for SMI | Primary care records | x | - |
| Medication for diabetes | Primary care records | x | - |
| *Process evaluation measures* |  |  |  |
| Mechanisms of Action questionnaire* | Self-report | - | x |
| Engagement with the Change One Thing app* | User data collected within the app | x | x |
| Acceptability of the intervention | Qualitative interviews | - | x |

^For logistical reasons it will not be possible to collect haemoglobin and cholesterol as part of this feasibility study. The feasibility of conducting blood tests is being tested through HbA1c assessment at baseline.
\*User data from the Change One Thing app will be collected throughout the intervention period while participants are engaging with the app. Information about Mechanisms of Action will be requested from participants via the app on a monthly basis and also collected at follow-up.

Figure E.1 (Appendix E) summarises the schedule of enrolment and recruitment, intervention delivery, and outcome assessments for this feasibility study.

#### Data collection

To assess the primary outcomes of this feasibility study (recruitment rate, attrition rate, and intervention delivery rate), the number of individuals approached, completing the intervention and/or signposted to other services, and providing outcome data will be recorded (along with withdrawals and, where available, reasons for withdrawal). Trained R&D staff at participating trusts will support participants to complete quantitative self-report measures as baseline and take physical measurements. Results will be recorded on standardised case report forms. Blood samples will be analysed at a centralised laboratory, with HbA1c results returned to the study team.

#### Objective 1 and 2 analyses

Data will be summarised descriptively using means and standard deviations (or medians and interquartile ranges) for continuous data and the frequency and proportion of events for categorical data. The number of sessions attended, and length of sessions will also be summarised. A Consolidated Standards for Reporting Trials (CONSORT) diagram will be used to display the flow of participants through the study.

To assess feasibility of quantitative data collection methods, we will record the proportion of missing data from questionnaires and case report forms, including data missing from medical records. We will also record the proportion of participants declining any of the physical measures (height, weight, blood test) and/or the accelerometers. Further, information about any data lost due to processing issues (e.g., blood results or accelerometer data) will be reported.

### 2.8 Objective 3: Evaluation of acceptability and feasibility of the DIAMONDS intervention

We will use a parallel mixed-methods approach in which qualitative and quantitative data will be collected to address this objective.

#### Recruitment and data collection

Quantitative process data will include: 1) descriptive markers of intervention content and mode of delivery (face to face or via telephone; digital; workbook; blended; remote or in-person); 2) dosage and reach: number of completed sessions, characteristics of participants; 3) satisfaction with intervention (sessions completed, ‘Did-not-attend’, dropouts).The research team will conduct semi-structured in-depth interviews with participants, carers/significant others, and DIAMONDS Coaches to determine how recipients responded to and engaged with the intervention across different contexts (e.g. service settings, organisations) and to examine the extent to which the study processes (including Coach training and support) were acceptable and fit for purpose.

##### Participants

At the point of consenting to take part in the DIAMONDS study, participants will have indicated their willingness (or not) to be contacted about taking part in an interview. From the pool of participants who have given consent to be contacted for an interview, we will select a purposive sample of up to 15 participants to include a range of ages and genders as well as intervention completers (i.e., participants who have completed at least 50% of intervention sessions within 16 weeks) and non-completers (i.e., participants who have completed less than 50% of intervention sessions within 16 weeks). If possible, we will also use information about baseline health, comorbidities, and level of engagement with the app and workbook to inform sampling. We will work with the trust R&D teams and the DIAMONDS Coaches to identify suitable participants if necessary.

A separate consent process, including the option of verbal consent for interviews undertaken remotely, will be in place, allowing participants to make an informed decision to contribute to this part of the study. If any risk of self-harm or suicide is identified during an interview, the researcher conducting the interview will follow the DIAMONDS Self-harm/Suicide Risk Protocol.

Interviews will last approximately 45 minutes and will explore experiences of recruitment, study procedures, and acceptability of the intervention (including important elements of the intervention and delivery).

##### Carers

We will aim to conduct interviews with seven carers, with interviews lasting approximately 45 minutes. For the purposes of this study, informal carers are defined as unpaid carers who are not subject to working regulations and provide support to a dependent person who they have a social relationship with, such as a spouse, other relative, neighbour, friend or other non-kin. Care includes support with household chores or other practical errands, transport to doctors or social visits, social companionship, emotional guidance, or help with arranging professional care.[56]

Carers will be given information about the interviews and full informed consent will be taken (verbally if necessary) before the start of the interview. Participants and carers will be interviewed alone to enable separate accounts to be generated. However, patients and carers may commonly wish to be interviewed together and in these cases, interviews will be run in a dyadic fashion. While dyadic patient-carer interviews can impose limits on response, they can also maximise opportunities for participants to co-construct their narratives about their experiences of the intervention and offer similar benefits to focus groups.[57]

##### DIAMONDS Coaches

We will aim to interview between five and ten Coaches, aiming for interview participation from at least one Coach from each participating study site as well as a range of NHS bands. We will aim to also interview Coaches who left the role before the end of the intervention delivery phase to learn about reasons for their withdrawal. To avoid any potential coercion from employers or co-workers, the research team, who are independent of employers, will approach Coaches to participate in semi-structured interviews. Informed consent will be secured from Coaches before the start of the interview. Coach interviews are expected to last approximately 30 minutes and will explore questions around the experience of being trained as a DIAMONDS Coach and delivering the intervention to study participants. Feedback on study and data collection procedures will also be sought and engagement with Coach support (mentoring calls, “booster” training sessions) recorded.

#### Objective 3 analysis

All interview data will be audio recorded (where appropriate; if not, extensive notes will be taken), transcribed verbatim (by a contracted third party) and analysed thematically using a Framework approach.[58] This process will draw on the theoretical framework of acceptability (TFA) that offers a robust and evidence-based approach to defining and evaluating anticipated and experienced forms of intervention delivery from the perspective of users and facilitators.[59] An initial coding framework will be developed, and transcripts checked against the framework to ensure that there are no significant omissions. Codes in each interview will be examined across individual transcripts as well as across the entire data set and allocated to the framework. Using aspects of the constant comparative method of analysis, broader categories using linking codes will be developed across interviews. Analysis will be guided by drawing on the theoretical foundations of the intervention and the findings from systematic reviews about determinants of self-management behaviour in SMI.

### 2.9 Objective 4: Exploratory Economic Analysis

An exploratory economic analysis will be conducted to evaluate the feasibility of collecting resource use and health-related quality of life data for the economic evaluation in the future RCT. As part of baseline assessment, participants will complete a service use questionnaire recording their utilisation of health (primary and secondary care) and social care resources. This will be complemented by a review of medical records. We will assess the results from both techniques to inform future data collection in the RCT.

#### Objective 4 analysis

To determine if questions about healthcare use may need to be adapted, the questionnaire response rates and the rate of missing data at item-level will be evaluated. The accuracy of the responses and the response consistency within the same participant will be checked to ensure that the questionnaires and the wording can yield accurate and reliable results. Both resource use (including intervention costs) and health-related quality of life outcomes will be summarised descriptively.

### 2.10 Objective 5: Acceptability and feasibility of Continuous Glucose Monitors

We will embed (as part of a PhD study led by JVEB), an evaluation of the acceptability of wearing continuous glucose monitors (CGMs). While use of CGMs is becoming increasingly widespread to support the self-management of diabetes across a range of populations, there is no evidence about the feasibility and acceptability of using this technology among people with diabetes and comorbid SMI. To address this gap in the evidence, we are planning an embedded mixed-methods evaluation of the acceptability and feasibility of the Abbott Freestyle Libre Pro[60] sensor within the DIAMONDS feasibility study. This is a blinded glucose sensor that is worn for 14 days, does not require calibration and provides 96 glucose readings/day (i.e. up to 1344 glucose readings per sensor). Using unblinded glucose sensors was considered but concerns were raised over potential anxiety related to the glucose readings and it was decided to use a blinded sensor in the first instance.

#### Recruitment and data collection

All participants will be asked to wear a blinded continuous glucose monitor (Abbott Freestyle Libre Pro) following the 16-week DIAMONDS intervention period. Sensors will be affixed to the participant’s upper arm by appropriately trained staff at the participating study sites. Participants will be asked to leave the sensor in place for 14 days and then return it to the study team for data download and analysis. Participants will remain blinded to the data collected by the sensor throughout.

Subsequently, participants and their informal carers will be asked to participate in semi-structured interviews to discuss their experience of having the sensor fitted, wearing, and removing the sensor as well as any problems they might have experienced with it. The interviews will also provide a chance to collect information about participants’ and carers’ attitudes about using technology to support self-management of their health more generally, e.g. through the use of wearables, such as activity trackers, or fitness or diet apps. Both participants who agreed to wear a continuous glucose monitor and those who declined will be invited for an interview.

The interviews about the continuous glucose monitors and attitudes towards technology-supported self-management will be separate from the process evaluation interviews as described previously.

#### Objective 5 analysis

To quantitatively assess the acceptability of the continuous glucose monitors, we will record the proportion of participants agreeing to wear a sensor, the average wear-period, the proportion of participants completing at least ten days of wear and of those completing at least five days, the number of sensors not returned, and any adverse events or side effects reported. In addition, the data collected by the sensors will be used produce ambulatory glucose profiles that will be further shown to study participants to help explain their glucose profile. As per international guidance, time-in-range data will be saved, together with glycaemic variability (measured as the coefficient of variation) and hypoglycaemic exposure,[61] while also recording HbA1c values. Due to the small sample size, these data will be summarised descriptively without any formal statistical analysis.

The same methods for recording and transcribing will be used for the interviews about CGMs as described for the interviews relating to the acceptability of the intervention. Similarly, a framework analysis approach will also be used for the CGM interviews drawing on the TFA to guide analysis. The coding will further be informed by findings from a systematic review of the acceptability of continuous glucose monitors with the most salient points identified in collaboration with DIAMONDS Voice. The focus of the analysis will be exclusively on the experience of wearing a CGM and attitudes about technology-supported self-management irrespective of thoughts or feelings about the DIAMONDS study or intervention.

### 2.11 Objective 6: Development of the intervention fidelity framework

Assessing fidelity of a complex intervention such as the DIAMONDS intervention is important in order to increase confidence in the interpretation of study results.[62] With this in mind, we will adapt tools and content from previous self-management programmes for long-term conditions, and use discussions with the study team, early findings from the feasibility study, the Coach training materials, and the intervention specification to determine the key components to include in the intervention fidelity framework. The framework will be developed iteratively and feasibility tested, in preparation for the main DIAMONDS RCT.

The intervention fidelity framework will consist of elements to measure: (i) adherence (whether the content of the intervention sessions was delivered as it was designed, including BCTs); (ii) dose (number of sessions delivered); (iii) quality of delivery of intervention sessions (BCTs and the manner/behaviour [both prescribed and proscribed] in which the coach delivers the programme); (iv) duration of the programme sessions (have the sessions been delivered within the estimated time?); and (v) participant responsiveness to the DIAMONDS intervention (was the intervention understood and ‘received’ by participants?).

#### Objective 6 data collection and analysis

In order to develop and refine the intervention fidelity tools (i.e. Coach behaviour checklists) used to assess the fidelity of Coach delivery, we will evaluate a sample of up to 10% of sessions (1:1 and group). Remote observation methods (e.g. through audio or video recording coaching sessions on encrypted devices) will be used for 1:1 sessions. Direct observations of group sessions will be carried out, unless COVID-19 restrictions prevent this; in which case they will be audio recorded. Trained assessors will use the intervention fidelity tools while observing/listening to the sample sessions. Three assessors will evaluate the recordings, check for consistency in scores and refine the tools where necessary until agreement is achieved so the tools are ready for use in a future RCT. Method of evaluation (e.g. direct observation or audio recordings) will also be documented to find out what is feasible for the main trial.

Refinement of the Coach training and Coach manual will be informed by findings from the qualitative interviews with coaches and participants (see objective 3 for further details); evaluation (pre- and post-) questionnaires of training (content, satisfaction, confidence with their role as a Coach); and group feedback with the Coaches post-training (sharing experience of the training and resources). Participant receipt will be measured via process evaluation / qualitative interviews. This will include specific determinants of self-management (e.g. participant engagement in writing their own action plan; cognitive strategies and behavioural skills used). This will enhance fidelity for the RCT.

### 2.12 Progression Criteria

Progression from the feasibility study to the RCT will be contingent on achieving:

1. Recruitment of at least 20 participants with Red-Amber-Green stop-go ratings based on <20 participants (Red); 20-25 (Amber); 26-30 (Green).
2. At least 50% of the intervention sessions delivered to 80% of participants.
3. Development of a coach training package.
4. Development of an intervention fidelity framework.
5. A self-management intervention that is acceptable and feasible to patients and Coaches. Any necessary modifications identified will be made prior to progression to a definitive trial.

## 3 Discussion

There is ample evidence showing that people with SMI experience substantial health inequalities that result in greater ill health, poorer quality of life, and reduced life expectancy than the general population. This so-called mortality gap is mainly driven by excess deaths among people with SMI attributable to preventable physical LTCs. The causes are complex and relate to behavioural factors (e.g. people with SMI are less likely to be non-smokers), social determinants such as unemployment and poor housing, the metabolic side effects of antipsychotic medication, and system level factors associated with disjointed care between physical and mental health teams, as well as primary and secondary care, and a lack of access to bespoke support.[44, 63, 64] However, there is now an emerging consensus that tackling excess mortality among people with SMI is a public health priority that demands interventions at the individual, system, and community level.[65]

We have mapped the mechanisms of action of health behaviours associated with physical health using the novel MoA framework to prompt identification of candidate behaviour change techniques with empirical evidence of effectiveness for managing LTCs.[43] Building on these findings and working with service users, carers, and healthcare professionals, we have developed an intervention to support people with SMI to self-manage type 2 diabetes. Before proceeding to test the clinical and cost effectiveness of our novel intervention, it is imperative that we first test the feasibility and acceptability of study procedures and intervention content and delivery. The main DIAMONDS RCT, which is planned to open for recruitment in 2022, will be one of the largest trials including people with SMI nationwide with a recruitment target of over 400 participants.

Research aimed at reducing health inequalities and the mortality gap is often considered challenging. However, we have previously shown that people with SMI are willing to participate in research and behavioural interventions have met with some success in reducing risk behaviours associated with poor physical health and premature mortality.[51] The DIAMONDS programme builds on this previous learning and maintains a critical focus on developing interventions to improve the physical health of people with SMI.

Furthermore, this feasibility study will also provide a platform to test out novel and innovative self-monitoring health technologies in the context of SMI and diabetes. There is a body of evidence to suggest that people with SMI engage differently with healthcare systems than the general population.[66-70] As such, it is important that they are included in research from an early stage to make sure they are not left behind in clinical and research innovations which could lead to a widening of the health inequalities already faced by this group.[71-73] To our knowledge, our study will be the first time that people with SMI have been invited to wear a continuous glucose monitor and share their feedback about their utility and acceptability.

### Dissemination plan

As part of our dissemination plan for this feasibility study, we will work with DIAMONDS Voice, our service-user and carer group, to communicate key milestones on social media and our project website. Where appropriate, DIAMONDS Voice members will be involved in the production and presentation of academic outputs, such as journal publications and conference submissions. We will continue to share findings and progress with our regional, national, and international networks through the DIAMONDS newsletter.

### COVID-19 contingency planning

The COVID-19 pandemic has reduced capacity in the NHS to support non-COVID clinical research, limiting the scope of this feasibility study. The single-group design preserves opportunities to test intervention feasibility and acceptability as well as feasibility of study procedures with reduced burden on the NHS. Strong evidence supports the feasibility of recruiting and randomising individuals with SMI and LTCs to trials of complex interventions (STEPWISE[52], SCIMITAR+[51]), offering crucial intelligence about the optimal ways to recruit to target in the context of randomised controlled trials for people with SMI. Additionally, the definitive DIAMONDS trial will include an internal pilot phase which will offer the means to check and refine strategies to recruit to target.

## Data Availability

N/A - Study protocol, no findings reported

BCT: behaviour change technique
BMI: Body Mass Index
CGM: continuous glucose monitoring
CTC: consent-to-contact
GP: general practitioner
HbA1c: glycated haemoglobin A1c
LTC: long-term condition
MoA: Mechanism of Action
NHS: National Health Service
NIHR: National Institute for Health Research
RCT: randomised controlled trial
R&D: research and development
SMI: severe mental illness
TFA: Theoretical Framework of Acceptability

## Acknowledgments

On behalf of the DIAMONDS Research Team we would like to thank DIAMONDS Voice, our service user and carer group, for their continued, enthusiastic, and expert contribution to the DIAMONDS Research Programme and specifically the participant-facing documents to be used in this study. We would also like to thank the NIHR Clinical Research Network Yorkshire & Humber for their continued support. Furthermore, we would like to thank our partners within the NHS for agreeing to be part of this study.

