## Appendices for "The DIAMONDS intervention to support self-management of type 2 diabetes in people with severe mental illness: study protocol for a single-group feasibility study"

### Appendix A: SPIRIT Checklist

| Section/item | Item No | Description | Addressed on page number |
| --- | --- | --- | --- |
| **Administrative information** | | |  |
| Title | 1 | Descriptive title identifying the study design, population, interventions, and, if applicable, trial acronym | 1 |
| Trial registration | 2a | Trial identifier and registry name. If not yet registered, name of intended registry | 3 |
|  | 2b | All items from the World Health Organization Trial Registration Data Set | N/A – not registered with WHO |
| Protocol version | 3 | Date and version identifier | N/A |
| Funding | 4 | Sources and types of financial, material, and other support | 22 |
| Roles and responsibilities | 5a | Names, affiliations, and roles of protocol contributors | Names & affiliations: pg 1&2; contributions: Author statement file included with submission |
|  | 5b | Name and contact information for the trial sponsor | Appendix C |
|  | 5c | Role of study sponsor and funders, if any, in study design; collection, management, analysis, and interpretation of data; writing of the report; and the decision to submit the report for publication, including whether they will have ultimate authority over any of these activities | Funder: pg 22; Sponsor: Appendix C |
|  | 5d | Composition, roles, and responsibilities of the coordinating centre, steering committee, endpoint adjudication committee, data management team, and other individuals or groups overseeing the trial, if applicable (see Item 21a for data monitoring committee) | Appendix C |
| Introduction |  |  |  |
| Background and rationale | 6a | Description of research question and justification for undertaking the trial, including summary of relevant studies (published and unpublished) examining benefits and harms for each intervention | 4 – 6 |
|  | 6b | Explanation for choice of comparators | N/A – single-group study |
| Objectives | 7 | Specific objectives or hypotheses | 6 |
| Trial design | 8 | Description of trial design including type of trial (eg, parallel group, crossover, factorial, single group), allocation ratio, and framework (eg, superiority, equivalence, noninferiority, exploratory) | 7 |
| Methods: Participants, interventions, and outcomes | | |  |
| Study setting | 9 | Description of study settings (eg, community clinic, academic hospital) and list of countries where data will be collected. Reference to where list of study sites can be obtained | 7&8 |
| Eligibility criteria | 10 | Inclusion and exclusion criteria for participants. If applicable, eligibility criteria for study centres and individuals who will perform the interventions (eg, surgeons, psychotherapists) | 7 |
| Interventions | 11a | Interventions for each group with sufficient detail to allow replication, including how and when they will be administered | 9-11 |
|  | 11b | Criteria for discontinuing or modifying allocated interventions for a given trial participant (eg, drug dose change in response to harms, participant request, or improving/worsening disease) | N/A – Feasibility study |
|  | 11c | Strategies to improve adherence to intervention protocols, and any procedures for monitoring adherence (eg, drug tablet return, laboratory tests) | N/A – Feasibility study |
|  | 11d | Relevant concomitant care and interventions that are permitted or prohibited during the trial | 6-11 |
| Outcomes | 12 | Primary, secondary, and other outcomes, including the specific measurement variable (eg, systolic blood pressure), analysis metric (eg, change from baseline, final value, time to event), method of aggregation (eg, median, proportion), and time point for each outcome. Explanation of the clinical relevance of chosen efficacy and harm outcomes is strongly recommended | Table 1; pg 12-14 |
| Participant timeline | 13 | Time schedule of enrolment, interventions (including any run-ins and washouts), assessments, and visits for participants. A schematic diagram is highly recommended (see Figure) | Appendix E |
| Sample size | 14 | Estimated number of participants needed to achieve study objectives and how it was determined, including clinical and statistical assumptions supporting any sample size calculations | 7 |
| Recruitment | 15 | Strategies for achieving adequate participant enrolment to reach target sample size | 7-9 |
| **Methods: Assignment of interventions (for controlled trials)** | | |  |
| Allocation: |  |  |  |
| Sequence generation | 16a | Method of generating the allocation sequence (eg, computer-generated random numbers), and list of any factors for stratification. To reduce predictability of a random sequence, details of any planned restriction (eg, blocking) should be provided in a separate document that is unavailable to those who enrol participants or assign interventions | N/A |
| Allocation concealment mechanism | 16b | Mechanism of implementing the allocation sequence (eg, central telephone; sequentially numbered, opaque, sealed envelopes), describing any steps to conceal the sequence until interventions are assigned | N/A |
| Implementation | 16c | Who will generate the allocation sequence, who will enrol participants, and who will assign participants to interventions | N/A |
| Blinding (masking) | 17a | Who will be blinded after assignment to interventions (eg, trial participants, care providers, outcome assessors, data analysts), and how | N/A |
|  | 17b | If blinded, circumstances under which unblinding is permissible, and procedure for revealing a participant’s allocated intervention during the trial | N/A |
| **Methods: Data collection, management, and analysis** | | |  |
| Data collection methods | 18a | Plans for assessment and collection of outcome, baseline, and other trial data, including any related processes to promote data quality (eg, duplicate measurements, training of assessors) and a description of study instruments (eg, questionnaires, laboratory tests) along with their reliability and validity, if known. Reference to where data collection forms can be found, if not in the protocol | 11-19 |
|  | 18b | Plans to promote participant retention and complete follow-up, including list of any outcome data to be collected for participants who discontinue or deviate from intervention protocols | N/A – (no follow-up) 14-15 |
| Data management | 19 | Plans for data entry, coding, security, and storage, including any related processes to promote data quality (eg, double data entry; range checks for data values). Reference to where details of data management procedures can be found, if not in the protocol | Appendix C |
| Statistical methods | 20a | Statistical methods for analysing primary and secondary outcomes. Reference to where other details of the statistical analysis plan can be found, if not in the protocol | 11-19 |
|  | 20b | Methods for any additional analyses (eg, subgroup and adjusted analyses) | N/A |
|  | 20c | Definition of analysis population relating to protocol non-adherence (eg, as randomised analysis), and any statistical methods to handle missing data (eg, multiple imputation) | N/A – feasibility study |
| **Methods: Monitoring** | | |  |
| Data monitoring | 21a | Composition of data monitoring committee (DMC); summary of its role and reporting structure; statement of whether it is independent from the sponsor and competing interests; and reference to where further details about its charter can be found, if not in the protocol. Alternatively, an explanation of why a DMC is not needed | Appendix C |
|  | 21b | Description of any interim analyses and stopping guidelines, including who will have access to these interim results and make the final decision to terminate the trial | N/A – Feasibility study |
| Harms | 22 | Plans for collecting, assessing, reporting, and managing solicited and spontaneously reported adverse events and other unintended effects of trial interventions or trial conduct | Appendix C |
| Auditing | 23 | Frequency and procedures for auditing trial conduct, if any, and whether the process will be independent from investigators and the sponsor | Appendix C |
| Ethics and dissemination | | |  |
| Research ethics approval | 24 | Plans for seeking research ethics committee/institutional review board (REC/IRB) approval | 6 |
| Protocol amendments | 25 | Plans for communicating important protocol modifications (eg, changes to eligibility criteria, outcomes, analyses) to relevant parties (eg, investigators, REC/IRBs, trial participants, trial registries, journals, regulators) | Appendix C |
| Consent or assent | 26a | Who will obtain informed consent or assent from potential trial participants or authorised surrogates, and how (see Item 32) | 8-9 |
|  | 26b | Additional consent provisions for collection and use of participant data and biological specimens in ancillary studies, if applicable | N/A |
| Confidentiality | 27 | How personal information about potential and enrolled participants will be collected, shared, and maintained in order to protect confidentiality before, during, and after the trial | Appendix C |
| Declaration of interests | 28 | Financial and other competing interests for principal investigators for the overall trial and each study site | Provided separately as part of submission to journal |
| Access to data | 29 | Statement of who will have access to the final trial dataset, and disclosure of contractual agreements that limit such access for investigators | Appendix C |
| Ancillary and post-trial care | 30 | Provisions, if any, for ancillary and post-trial care, and for compensation to those who suffer harm from trial participation | N/A |
| Dissemination policy | 31a | Plans for investigators and sponsor to communicate trial results to participants, healthcare professionals, the public, and other relevant groups (eg, via publication, reporting in results databases, or other data sharing arrangements), including any publication restrictions | 21 |
|  | 31b | Authorship eligibility guidelines and any intended use of professional writers | 22 |
|  | 31c | Plans, if any, for granting public access to the full protocol, participant-level dataset, and statistical code | Appendix C |
| Appendices |  |  |  |
| Informed consent materials | 32 | Model consent form and other related documentation given to participants and authorised surrogates | Appendix B |
| Biological specimens | 33 | Plans for collection, laboratory evaluation, and storage of biological specimens for genetic or molecular analysis in the current trial and for future use in ancillary studies, if applicable | N/A |

SPIRIT 2013 Checklist: Recommended items to address in a clinical trial protocol and related documents*

*It is strongly recommended that this checklist be read in conjunction with the SPIRIT 2013 Explanation & Elaboration for important clarification on the items. Amendments to the protocol should be tracked and dated. The SPIRIT checklist is copyrighted by the SPIRIT Group under the Creative Commons “[Attribution-NonCommercial-NoDerivs 3.0 Unported](http://www.creativecommons.org/licenses/by-nc-nd/3.0/)” license.

### Appendix B: Sample consent form

**PARTICIPANT CONSENT FORM**

**Title of Study: DIAMONDS**

**IRAS ID: 279019**

**REC Reference: Leeds West REC 21/YH/0059**

**Participant ID:**

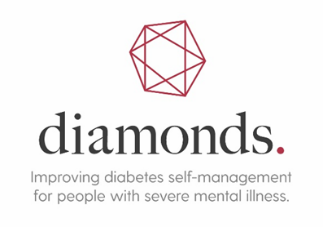

If you wish to take part in the DIAMONDS study, **please place your initials in each of the boxes below, sign and date this form.**

*Please* ***initial*** *each box*

***Initials***

| 1. I confirm that I have read and understand the information sheet version [xx] dated [xxxx] for the above study and have had the opportunity to ask any questions about the study and any questions have been answered to my satisfaction.   ***Initials*** | | | | | | | | | | | | | | | | | | | | | | | | |
| --- | --- | --- | --- | --- | --- | --- | --- | --- | --- | --- | --- | --- | --- | --- | --- | --- | --- | --- | --- | --- | --- | --- | --- | --- |
| 2. I understand that my participation is voluntary and that I am free to withdraw at any time without giving any reason, and without my medical care or legal rights being affected.   ***Initials***   3. I agree to the University of York holding copies of my consent form, other study related documents and my contact details to allow them to contact me for appointments and send me letters and questionnaires.   ***Initials***   4. I agree to my GP being informed of my participation in the study and being advised of any significant information relating to my health that comes to light during my participation. 5. I understand that relevant sections of my hospital/GP medical notes and data collected during the study, may be looked at by individuals from the University of York, from regulatory authorities or from the NHS Trust, where it is relevant to my taking part in this research. I give permission for these individuals to have access to my records.   ***Initials***  ***Initials***   6. I understand that the information collected about me could be used to support other research in the future, and may be shared anonymously with other researchers. 7. I understand that the information held and maintained by [*enter name of organisation(s) that will provide the data*] and other central UK NHS bodies may be used to help contact me or provide information about my health status.   ***Initials***  ***Initials***   8. **I agree to take part** **in the DIAMONDS Study.** | | | | | | | | | | | | | | | | | | | | | | | | |
| In addition to the above statements please initial the following boxes to indicate whether you agree with the following statements. Your participation in this research study will not be affected if you do not agree with these statements.  **Yes No**  ***Initials***  ***Initials***  I am happy to be contacted about giving feedback on the  DIAMONDS study  ***Initials***  ***Initials***  I would like to receive a summary of the study findings | | | | | | | | | | | | | | | | | | | | | | | | |
|  | |  | *d* | | *d* | | */* | | *m* | | *m* | | */* | | *y* | | *y* | | *y* | | *y* | |  | |
| Print name |  | | |  | |  | | / |  |  | | / | | 2 | | 0 | |  | |  | |  | | Signature |
| Name of participant (*please print*) | | | | Date | | | | | | | | | | | | | | | | | | Signature of participant | | |
|  | | | | *d* | | *d* | | */* | *m* | *m* | | */* | | *y* | | *y* | | *y* | | *y* | |  | | |
| Print name |  | | |  | |  | | / |  |  | | / | | 2 | | 0 | |  | |  | |  | | Signature |
| Name of person taking consent  (*please print*) | | | | Date | | | | | | | | | | | | | | | | | | Signature of person taking consent | | |

**[Original to be kept in Trial Master File; one copy given to participant; one copy sent to participant’s GP]**

### Appendix C: Study Management and Governance Structures

#### Study sponsorship

Bradford District Care NHS Foundation Trust will act as the lead organisation and contractual partner with the NIHR and as such will hold overall responsibility for the delivery of the programme. The University of York will act as sponsor for this feasibility study.

#### The DIAMONDS Programme Management Team

This feasibility study forms part of the DIAMONDS research programme which is led by Professor of Psychiatry (Chief-investigator). The day-to-day running of the programme, including the feasibility study, is overseen by the Programme Manager with support from the workstream leads who are experts in health services research methods and diabetes, respectively. Statistical oversight is provided by a Professor of Trials and Statistics.

Decisions about the feasibility study and the programme as a whole are made by the Programme Management Team which includes experts in diabetes, mental health, statistics, and health economics, as well as GPs, psychologists, and service user and carer representatives.

#### The DIAMONDS Programme Steering Committee

The Programme Management Team is accountable to and supported by the Programme Steering Committee which also fulfils the role of the Data and Ethics Committee (DMEC) for the feasibility study. The Steering Committee membership consists of an academic GP, a trialist, a statistician, a psychiatrist, a diabetologist, and a service user representative.

#### Monitoring adverse events

##### Definitions

An **adverse event** is any unexpected effect or untoward clinical event affecting the participant. It can be directly related, possibly related or completely unrelated to the intervention. It can also be classed according to severity, such that non-serious Adverse Event (AE) includes discomfort or slight worsening of symptoms, or Serious Adverse Event (SAE), which may be particularly harmful, dangerous or require hospitalisation.

Hospitalisations for treatment planned prior to enrolment and hospitalisation for elective treatment of a pre-existing condition will not be considered as an SAE. Complications occurring during such hospitalisation will be AEs.

##### Detecting and recording AEs and SAEs

Any AEs or SAEs will be reported to the Chief Investigator and will be reviewed by a clinician independent to the DIAMONDS study team. This will include all reported cases of COVID-19. The reporting period will be from study entry up to the last follow-up visit. Details about AEs/SAEs will be captured at each contact with a DIAMONDS Coach or study assessment. AEs/SAEs that might have occurred since the previous visit or assessment are elicited from the patient by open questioning and recorded. All events related to the DIAMONDS intervention will be recorded on adverse events forms. Further information may be requested for follow up of these events. Detailed records will be kept of all adverse events.

##### Evaluation of AEs and SAEs

Adverse events that are deemed possibly, probably or definitely related to participation in this study and all SAEs will be evaluated for seriousness, causality, severity and expectedness by the chief investigator and reviewed by an independent clinician/mental health specialist. All AEs/SAEs will be reviewed in terms of suspected causal relationship (e.g. unrelated, unlikely, possibly, probably, definitely) to the study intervention.

##### Reporting AEs and SAEs

All SAEs will be reported to the sponsor and to the Research Ethics Committee (REC) in line with their guidelines. Serious events deemed unexpected and related events will usually be reported to the REC within 15 days of the event being reported. All others will be reported in the usual 6-monthly progress report. Any relevant further information will be subsequently communicated, and events will be followed up until the event is resolved or a decision is made that no further follow-up is necessary. In addition, all associated investigators will be notified. The numbers and details of all AEs and SAEs will be reported to the PMT and PSC.

AEs reported by study participants that are not classified as an SAE will be reported and included in reports submitted to the PSC in agreement with the committee chair.

Where repeated adverse events (serious or non-serious) of a similar type are observed, these will be discussed with the PMT and other relevant groups and will be onward reported to the REC and Sponsor should concerns be raised in relation to the type of event and/or frequency observed.

#### Suicide and self-harm risk management

Inherent in the population under scrutiny is the risk of self-harm and suicide. We will follow good clinical practice and follow a Risk Protocol for the monitoring of suicide and self-harm risk during all encounters with study participants. The study team have a wealth of experience in developing and implementing risk protocols for use in studies involving psychological interventions for SMI. Where any risk to participants, due to expressed thoughts of self-harm or suicide, is encountered, a risk assessment will be conducted. Level of risk will be determined and discussed with a clinical member of the study team. Risk will be reported to the participant’s GP (with the participant’s consent) where deemed necessary by a clinical member of the research team. Acute risk will be dealt with immediately and will involve a clinical member of the DIAMONDS study team. At least one clinical member of the team will be on call at all times in order to respond to risk whilst participants are involved in the study. All members of the study team as well as DIAMONDS Coaches and members of R&D teams involved in data collection at participating trusts will complete training on the risk protocol, delivered by a clinical member of the team, before commencing contact with participants. Members of the study team, DIAMONDS Coaches, and R&D staff will be provided with support and debriefing following risk if required.

#### Duty of care

We will use YTU standard operating procedures to support researchers, DIAMONDS Coaches, and R&D staff to report to GPs or responsible services instances where there are concerns about the health of the participant.

A distress protocol has been developed and will be followed in case any participants displays signs of distress during the qualitative interviews. Participant wellbeing will be the main priority and researchers conducting the interviews will sign-post further support as needed.

#### Researcher safety and lone working

Researcher safety is of paramount importance. We will use the York Trials Unit (YTU) standard operating procedures for fieldwork and lone working (see Appendix 1). Fieldwork is defined as any research activity that involves data collection either on-site (university premises) or off-site (e.g. patient’s homes, hospitals premises, and community centres). All researchers tasked with fieldwork will undertake lone worker training and conduct a risk assessment with their line manager about the specific tasks to be carried out. Researchers will be able to appoint a designated person (academic or administrative staff) who will act as a safety contact. A system will be agreed between the researcher and the designated person to communicate when the fieldwork trip has started and finished. Researchers will have regular debriefs with their line manager and the Chief Investigator to review this process and check that it is fit for purpose. Details of the lone worker policy will be included in the researcher handbook.

#### Statement of indemnity and complaint handling

Normal NHS indemnity procedures will apply. The University of York will also provide relevant cover. The PIS will provide participants with contact details of the Sponsor in case of complaint. If there is negligent harm during the study, when the NHS Trust owes a duty of care to the person harmed, NHS indemnity covers NHS staff and medical academic staff with honorary contracts only when the feasibility study has been approved by the R&D department. NHS indemnity does not offer no-fault compensation and is unable to agree in advance to pay compensation for non-negligent harm.

#### Monitoring quality control and assurance

Quality control will be maintained through adherence to Standard Operating Procedures (SOPs), study protocol, the principles of ICH/GCP, research governance and relevant study regulations. The intervention delivered in this feasibility study is low risk and major safety data are not anticipated. Monitoring of study conduct and data collected will be performed by a combination of central review and site monitoring visits to ensure the study is conducted in accordance with good clinical practice. The main areas of focus will include consent, serious adverse events, and essential study documents. All monitoring findings will be reported and followed up with the appropriate persons in a timely manner. The study may be subject to inspection and audit by the University of York under their remit as sponsor and other regulatory bodies to ensure adherence to GCP. The investigator(s)/institutions will permit study-related monitoring, audits, REC review and regulatory inspection(s), providing direct access to source data/documents.

Data collected as part of this research includes questionnaires, clinical assessments, information from medical records, and qualitative data from interviews. Data will be collected through designed questionnaires on paper. These paper forms will be scanned at YTU and the data stored in a database where they are checked against the hard copy of the questionnaire. Data is error checked and validation checks are run against the database. Discrepancies identified during validation which require resolution are communicated to the relevant person who is in a position to obtain the information required to rectify the discrepancy. If data are found to be missing from participant completed questionnaires, participants will be contacted by one of the research team members in an attempt to collect the data.

DIAMONDS Coaches will have the option to record intervention logs electronically via a secure website.

#### Data management

In line with the 2018 General Data Protection Regulation and the UK Policy Framework for Health and Social Care,^60^ anonymised feasibility study data will be securely archived by the University of York for a minimum of 10 years. Personal data of participants will be stored for up to three years after the study has ended for the purpose of disseminating study findings. It is unlikely that this will take longer than 12 months, however, to ensure that participants receive adequate and full information about the study after it has finished, additional time has been allocated.

All information collected during the feasibility study will be kept strictly confidential as detailed above. Information will be held securely in paper and/or electronic formats at the University of York. The University of York complies with all aspects of the 2018 General Data Protection Regulation and Data Protection Act 2018. Operationally this will include obtaining explicit consent from study participants to record personal details including name, postal and email address, and contact telephone numbers, and appropriate storage, restricted access and disposal arrangements for their personal details. All participants will be informed of their rights in regard to the personal information stored, including erasure, rectification and objection. All work will be conducted following the University of York’s data protection guidance which is publicly available (University of York, 2018).^61^

A data protection impact assessment (DPIA) has been completed and approved by the University of York’s data protection team. In line with University of York policy, this will be kept under review throughout the duration of the study.

#### Confidentiality

At recruitment, each participant will be allocated a unique study identification number. This number will be used to identify participants throughout the study. The master register linking participants personal and contact details with the identifier will be maintained by the York Trials Unit data manager. Only relevant members of the study team will have access to this information via a password protected database within secure offices. A Participant Screening/Enrolment Log will be maintained, providing the dates patients were screened, whether they were eligible or not (with reason) and if consented or not (with reason). This log will not contain any identifiable patient details.

Clinical information will not be released without the written permission of the participant, except as necessary for monitoring and auditing by the Sponsor, its designee, Regulatory Authorities, or the REC. The investigator and study site staff involved with this study may not disclose or use for any purpose other than performance of the study, any data, record, or other unpublished, confidential information disclosed to those individuals for the purpose of the study. Prior written agreement from the Sponsor or its designee must be obtained for the disclosure of any said confidential information to other parties.

#### Data security

- All data will be stored in accordance with data protection requirements and will be kept either in a locked filing cabinet in a secure office or in the case of electronic data on a secure sever with a password protected computer and files.
- Personal addresses, postcodes and other contact details of consenting participants will be stored on a secure password-protected server located at the University of York, for the purposes of assisting in follow-ups during the study. All personally identifiable participant data will be coded, pseudonymised by participant number in all manual and electronic files. YTU will maintain a list of participant identification numbers for all study participants at each site.
- Interview recordings will be downloaded onto a password protected computer and deleted from the recording device. They will then be securely uploaded to a GDPR-compliant transcribing company.
- No data will be stored on a home computer or laptop.
- All data will be stored for a maximum of 10 years, which will allow time for any academic challenge to be made. All data personal will be deleted after this time.

### Appendix D: TIDieR Checklist

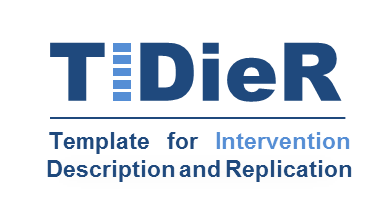

**The TIDieR (Template for Intervention Description and Replication) Checklist*:**

Information to include when describing an intervention and the location of the information

| **Item number** | **Item** | **Where located **** | |
| --- | --- | --- | --- |
|  |  | Primary paper  (page or appendix  number) | Other ^†^ (details) |
|  | **BRIEF NAME** |  |  |
| **1.** | Provide the name or a phrase that describes the intervention. | 9 | ______________ |
|  | **WHY** | 9-11 |  |
| **2.** | Describe any rationale, theory, or goal of the elements essential to the intervention. |  | _____________ |
|  | **WHAT** |  |  |
| **3.** | Materials: Describe any physical or informational materials used in the intervention, including those provided to participants or used in intervention delivery or in training of intervention providers. Provide information on where the materials can be accessed (e.g. online appendix, URL). | 9-11 | _____________ |
| **4.** | Procedures: Describe each of the procedures, activities, and/or processes used in the intervention, including any enabling or support activities. | 9-11 | _____________ |
|  | **WHO PROVIDED** |  |  |
| **5.** | For each category of intervention provider (e.g. psychologist, nursing assistant), describe their expertise, background and any specific training given. | 9 | _____________ |
|  | **HOW** |  |  |
| **6.** | Describe the modes of delivery (e.g. face-to-face or by some other mechanism, such as internet or telephone) of the intervention and whether it was provided individually or in a group. | 9-11 | _____________ |
|  | **WHERE** |  |  |
| **7.** | Describe the type(s) of location(s) where the intervention occurred, including any necessary infrastructure or relevant features. | 10-11 | _____________ |
|  | **WHEN and HOW MUCH** |  |  |
| **8.** | Describe the number of times the intervention was delivered and over what period of time including the number of sessions, their schedule, and their duration, intensity or dose. | 9-11 | _____________ |
|  | **TAILORING** |  |  |
| **9.** | If the intervention was planned to be personalised, titrated or adapted, then describe what, why, when, and how. | 9-11 | _____________ |
|  | **MODIFICATIONS** |  |  |
| **10.^ǂ^** | If the intervention was modified during the course of the study, describe the changes (what, why, when, and how). | N/A | Feasibility study protocol |
|  | **HOW WELL** |  |  |
| **11.** | Planned: If intervention adherence or fidelity was assessed, describe how and by whom, and if any strategies were used to maintain or improve fidelity, describe them. | 18-20 | _____________ |
| **12.^ǂ^** | Actual: If intervention adherence or fidelity was assessed, describe the extent to which the intervention was delivered as planned. | N/A | _Feasibility study protocol |

** **Authors** - use N/A if an item is not applicable for the intervention being described. **Reviewers** – use ‘?’ if information about the element is not reported/not sufficiently reported.

† If the information is not provided in the primary paper, give details of where this information is available. This may include locations such as a published protocol or other published papers (provide citation details) or a website (provide the URL).

ǂ If completing the TIDieR checklist for a protocol, these items are not relevant to the protocol and cannot be described until the study is complete.

* We strongly recommend using this checklist in conjunction with the TIDieR guide (see *BMJ* 2014;348:g1687) which contains an explanation and elaboration for each item.

* The focus of TIDieR is on reporting details of the intervention elements (and where relevant, comparison elements) of a study. Other elements and methodological features of studies are covered by other reporting statements and checklists and have not been duplicated as part of the TIDieR checklist. When a **randomised trial** is being reported, the TIDieR checklist should be used in conjunction with the CONSORT statement (see [www.consort-statement.org](http://www.consort-statement.org)) as an extension of **Item 5 of the CONSORT 2010 Statement.** When a **clinical trial** **protocol** is being reported, the TIDieR checklist should be used in conjunction with the SPIRIT statement as an extension of **Item 11 of the SPIRIT 2013**

### Appendix E: Spirit Figure

|  | **STUDY PERIOD** | | | | | | | |
| --- | --- | --- | --- | --- | --- | --- | --- | --- |
|  | **Enrolment** | **Post-enrolment** | | | | | **Close-out** |  |
| **TIMEPOINT**** | ***-t_1_*** | ***t_1_*** | ***t_2_*** | ***t_3_*** | ***…*** | ***t_16_*** | ***t_x_*** | ***t_x+1_*** |
| **ENROLMENT:** |  |  |  |  |  |  |  |  |
| **Eligibility screen** | X |  |  |  |  |  |  |  |
| **Informed consent** | X |  |  |  |  |  |  |  |
| **INTERVENTIONS:** |  |  |  |  |  |  |  |  |
| ***The DIAMONDS Intervention*** |  |  |  |  |  |  |  |  |
| **ASSESSMENTS:** |  |  |  |  |  |  |  |  |
| ***Demographics*** | X |  |  |  |  |  |  |  |
| ***Psychological health*** | X |  |  |  |  |  |  |  |
| ***Diabetes measures*** | X |  |  |  |  |  |  |  |
| ***Accelerometer*** |  |  |  |  |  |  |  |  |
| ***Health economic outcomes*** | X |  |  |  |  |  |  |  |
| ***Process evaluation measures**** | X |  |  |  |  |  | X |  |
| ***Qualitative interviews (intervention acceptability)*** |  |  |  |  |  |  | X |  |
| ***Continuous glucose monitor*** |  |  |  |  |  |  |  |  |
| ***Qualitative interviews (CGM)*** |  |  |  |  |  |  |  | X |

Figure E.1: Summary of the enrolment, intervention delivery, and data collection schedule for the DIAMONDS feasibility study.
